# Comparison of Decision Modeling Approaches for Health Technology and Policy Evaluation

**DOI:** 10.1101/2020.05.26.20113845

**Authors:** John Graves, Shawn Garbett, Zilu Zhou, Jonathan S. Schildcrout, Josh Peterson

**Affiliations:** Department of Health Policy, Vanderbilt University School of Medicine, Vanderbilt University Medical Center, 2525 West End Avenue, Suite 1200, Nashville, TN 37203; Vanderbilt University Medical Center, Department of Biostatistics; Health Policy Analyst II, Vanderbilt University Medical Center Department of Health Policy; Department of Biostatistics, Vanderbilt University School of Medicine, Vanderbilt University Medical Center; Vanderbilt University School of Medicine, Vanderbilt University Medical Center, Department of Informatics

**Keywords:** Decision modeling, pharmagogenomics, health economic methods

## Abstract

We discuss tradeoffs and errors associated with approaches to modeling health economic decisions. Through an application in pharmacogenomic (PGx) testing to guide drug selection for individuals with a genetic variant, we assessed model accuracy, optimal decisions and computation time for an identical decision scenario modeled four ways: using (1) coupled-time differential equations [DEQ]; (2) a cohort-based discrete-time state transition model [MARKOV]; (3) an individual discrete-time state transition microsimulation model [MICROSIM]; and (4) discrete event simulation [DES]. Relative to DEQ, the Net Monetary Benefit for PGx testing (vs. a reference strategy of no testing) based on MARKOV with rate-to-probability conversions using commonly used formulas resulted in different optimal decisions. MARKOV was nearly identical to DEQ when transition probabilities were embedded using a transition intensity matrix. Among stochastic models, DES model outputs converged to DEQ with substantially fewer simulated patients (1 million) vs. MICROSIM (1 billion). Overall, properly embedded Markov models provided the most favorable mix of accuracy and run-time, but introduced additional complexity for calculating cost and quality-adjusted life year outcomes due to the inclusion of “jumpover” states after proper embedding of transition probabilities. Among stochastic models, DES offered the most favorable mix of accuracy, reliability, and speed.

## 1. Introduction

Decision analytic assessments of health policies and technologies play an important role in determining the pricing and reimbursement of health care services worldwide. These assessments draw on a range of modeling methods to inform policy and clinical decision making. Commonly used methods include decision trees, cohort and individual state transition (Markov) models, discrete event simulation, stochastic process theory models based on differential equations, and hybrid models that blend elements across approaches (e.g., Discretely Integrated Condition Event models; see Brennan et al., 2006; Caro, 2016; Karnon, 2003; Rutter et al., 2011; Stahl, 2008; van Rosmalen et al., 2013).

For a given health technology assessment or research question, best practice recommends model choice based on consultation with clinical, methodological, and policy experts (Caro and Möller, 2016; Stahl, 2008). This ensures that the model represents essential elements of the underlying disease or therapeutic processes, as well as the alternative strategies under consideration.

In practice, considerations over model transparency, detail, ease of use, and even regulatory disclosure requirements, also may come into play. In addition, the application of value of information (VOI) methods may inform modeling choices due to the computational demands of VOI (Heath et al., 2019; Jalal et al., 2015; Jalal and Alarid-Escudero, 2017; Kunst et al., 2019). In other words, different modeling choices may be made depending on the weight placed on maximising the resolution and scope of output for a current decision problem on the one hand, and on understanding parameter uncertainty to refine future research priorities on the other.

Our objective for this study was to compare alternative approaches to modeling health economic decisions. Specifically, we assessed model outputs, optimal decisions and model computation time for an identical decision problem modeled four ways: using a discrete time cohort state transition (MARKOV) model, an individual discrete time state transition microsimulation (MICROSIM) model, discrete event simulation (DES), and a stochastic theory-based model based on time delay differential equations (DEQ). Importantly, these models drew on an identical set of underlying parameters that should, we hypothesized, lead to equivalent (in expectation) cost and quality-adjusted life year (QALY) estimates and decision outcomes. Our results demonstrate that using commonly-applied discrete time model structure and adjustment methods, they do not.

An important contribution of our study is the exposition of key sources of modeling error and how they can affect the accuracy, reliability and resolution of model outputs. We demonstrate through an application how these various errors can play out in practice. In particular, we show that adjustments must be made to discrete time individual and cohort state transition models to produce equivalent estimates as DES and DEQ models. These adjustments are necessary due to an interaction between competing events and the coarsening of continuous time into discrete time cycles—and are important in (common) situations where literature-based parameters are embedded within transition probability matrices in discrete-time Markov models. Finally, we also show that to achieve a given degree of output reliability, discrete time microsimulation models require considerably more Monte Carlo draws than a DES model. We include replication code and simulation functions for all model types considered.

## 2. Background

### 2.1 Modeling Types

The following section provides basic information on the model types we consider. This information is not intended to be exhaustive of the details and assumptions of each method; we leave such comprehensive treatment to the literature (Brennan et al., 2006; Drummond et al., 2015; Neumann et al., 2016). Rather, we highlight specific aspects of each modeling approach that are germane to issues of output scope, resolution, variance/bias, and computational efficiency that we highlight in our results.

#### 2.1.1 Discrete Time Cohort and Individual State Transition Models

State transition models (STM) are the most widely used modeling framework in health economic evaluation (Karnon, 2003). STMs are conceptualised around a set of mutually exclusive and collectively exhaustive states (e.g., healthy, sick, dead). Models are further defined by an initial occupancy vector (i.e., a count or fraction of patients in each state at baseline) and a set of state transition rates (for a continuous time model) or probabilities at a specified cycle length (for a discrete time model). The vast majority of health economic modeling applications are based on the discrete time case (van Rosmalen et al., 2013), so we focus on that model type here.

In a discrete-time STM, an initial occupancy vector is modeled forward using a transition probability matrix for a defined time horizon or until all individuals enter an absorbing state (e.g., death). Values, such as health-related quality of life and costs, are assigned to each state. These state values, along with information on the amount of person-time spent in each state, are summarized (often with discounting) to produce outcome estimates on various quantities of interest (e.g., expected life expectancy, expected quality-adjusted life expectancy, and expected costs).

A key decision when considering a STM is whether to jointly simulate the collective experience of a cohort, or to simulate individual patient trajectories via first order Monte Carlo microsimulation. Advantages of cohort STMs (often referred to as Markov models) include their simplicity, computational efficiency, and ease of use. A disadvantage is that the Markovian “memoryless” assumption complicates applications in which patient history affects transition probabilities (Siebert et al., 2012). While tunnel states provide a workaround, they can result in models with large numbers of states (i.e., “state explosion”; see Siebert et al., 2012).

Microsimulation models are useful in applications where state transition probabilities depend on a patient’s history (e.g., time since disease onset, the occurrence of previous events, or time-varying response to treatment; see Rutter et al., 2011). More broadly, microsimulation models facilitate transition probabilities that are a function of any number of attributes. Microsimulation also affords the flexibility to capture a greater scope of outputs since the model can return estimates of the entire distribution of events, rather than just expected values. This level of detail could be important for informing multi-criteria decision analyses that base decisions on a wider array of criteria beyond expected quality-adjusted life expectancy and costs (Baltussen et al., 2019; Baltussen and Niessen, 2006; Peacock et al., 2009; Thokala and Duenas, 2012).

The flexibility afforded by microsimulation comes at a cost, however, since the computational burden of microsimulation is large. As described below, these computational demands derive from three primary sources: first-order (stochastic) error; the need to simulate all time cycles for each individual, even if there is no event; and an additional increase in model output variance and bias that occurs when the precise timing of events are “truncated” to occur at cycle boundary points.

#### 2.1.2 Discrete Event Simulation

Discrete Event Simulation (DES) is a modeling methodology designed to incorporate the timing and interdependency of events (Caro and Möller, 2016; Karnon et al., 2012; Standfield et al., 2014). Though its origins are in industrial engineering and operations research, DES is increasingly used in health technology assessments (Jacobson et al., 2006; Karnon, 2003; Karnon et al., 2012; Stahl, 2008; Standfield et al., 2014).

DES models are similar to microsimulation models in that they simulate individual patient trajectories. As such, they can also be computationally demanding. One advantage, however, is that DES further extends the flexibilities of microsimulation. For example, in a DES model it is straightforward to allow the probability of some future event to depend on the time spent in a given state. In addition, DES models can more easily incorporate interdependencies in the timing of events and/or restrictions on available resources based on queuing or other constraints. Another advantage is that DES models the precise timing of events themselves (i.e., a DES model does not cycle through time periods when no events occur). These advantages afford DES considerably more modeling flexibility and computational efficiency as compared with microsimulation.

#### 2.1.3 Differential Equations Modeling

Many decision and disease processes can be represented using stochastic process theory to deterministically solve for key outcomes (van Rosmalen et al., 2013). These processes can be represented in terms of differential equations (DEQs), the solution to which provides the occupancy of an underlying process at any time *t*. Expected values of key outcomes (e.g., average discounted QALYs and costs) are obtained by integrating this solution from baseline (i.e., *t=0)* to some specified time *T*. Solutions to this integration problem have been developed for health economic applications in which transition rates are constant over time and/or evolve as a piecewise function of age (van Rosmalen et al., 2013). Standard differential equations can model a process that conforms to the Markovian “memoryless” property, while coupled time delay differential equations can model processes in which the solution at any time depends on previous values of the function (Graves et al., 2018).

Estimates derived from the numerical solution to a DEQ deterministically solve for expected outcomes, and thus eliminate stochastic uncertainty inherent to DES and microsimulation. Commonly available numerical solvers provide relatively fast solutions. A downside, however, is that the scope of modeled outputs is limited: the model solves for average outcomes but does not provide insights into higher order moments (e.g., variance). Moreover, specification of the model is challenging—and making even minor changes to model structure is a nontrivial task. DES and microsimulation methods, by comparison, return information on the entire distribution of events and can be easily constructed and modified without advanced mathematical training.

### 2.2 A Taxonomy of Errors in Health Economic Modeling

As discussed above, modeling choices affect computational demands and the range of outputs that can be analysed. Model structure also determines the types of error—that is, differences between quantities of interest based on the underlying event generation process and the model’s output—that must be accounted for. Some errors are well-known and are shared across modeling approaches. Others are more nuanced, less widely acknowledged, and specific to certain modeling strategies. Finally, depending on their size, errors may nor may not affect optimal decisions—though ex ante it is difficult to know which errors could affect decision-making for any given application. Thus, it is important that modeling practitioners be aware of sources of error, the situations in which they arise, and ways to address them.

This section, as well as Table 1, outlines a taxonomy of modeling error. We focus in particular on errors that can affect estimates and decisions, and further distinguish between **errors of reliability and uncertainty** (i.e., those that can be addressed by increasing the number of simulated patients or through further research) and **errors in expectation** (i.e., those that, even with an infinite number of simulated patients or with parameters with little-to-no uncertainty, would yield biased estimates that may result in different optimal decisions). We discuss ways to adapt model specification and execution to reduce and/or eliminate sources of error, and cover specific details on implementation of these corrections in the methods section below.

**Table 1.**
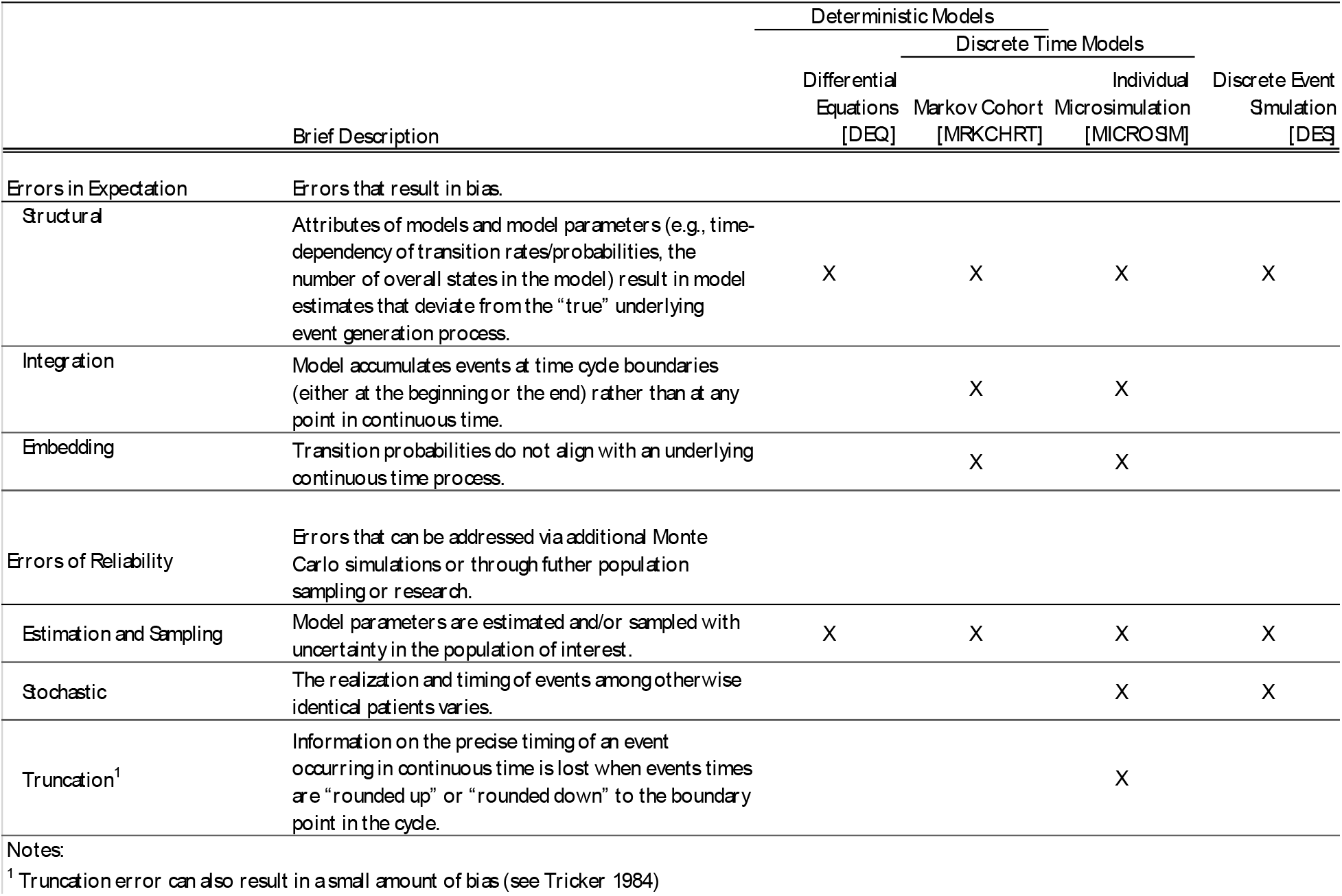
Typology of Errors

To understand our error taxonomy it is useful to conceptualise the modeled process (e.g., the progression of a disease) occurring in continuous time. Ideally, one has data on the precise timing of all events experienced by the population of interest. In that case, one can estimate all competing event rates directly using standard statistical approaches (e.g., using a multi-state model of event duration outcomes). Transitions among discrete states, moreover, can be summarized in terms of transition intensities in continuous time.

In practice, models must deviate from this ideal in meaningful ways. Researchers often do not have access to comprehensive data on the population of interest. Model parameters must therefore be curated from the published literature, or may be estimated via calibration with aggregate data on disease prevalence, incidence, etc. (Briggs, 2006). Literature-based parameters can take on any number of “flavors” (e.g., estimated event rates, odds ratios, probabilities, etc.) and there is no guarantee that what is published maps directly into the selected model structure. Therefore, parameters are often transformed (e.g., rates are converted into probabilities for the selected cycle length) so they can be embedded within the model.

Even when the underlying data or parameters are available, assumptions are often made over whether model parameters are fixed or evolve differentially over time and/or with specific patient attributes (e.g., race, age, gender, etc.). These questions are particularly germane to models that draw on literature-based parameters because the cohort and/or population data underlying these parameters often do not align with the population of interest.

We classify **structural error** as attributes of models and model parameters (e.g., time-dependency of transition rates/probabilities, the number of overall states in the model) that result in model estimates that deviate from the “true” underlying event generation process. It is worth differentiating structural errors from parameter heterogeneity, or differences in model parameters across relevant population sub-groups. In the case of heterogeneity, the overall structure of the model could accurately represent the event generation process. However, decisions could be improved by estimating model outputs and making separate decisions for each sub-population (Briggs et al., 2012).

Model parameters are also often subject to sampling and estimation uncertainty. **Estimation errors** are inherent to all model types discussed here. Understanding the role of estimation uncertainty is the focus of a growing body of research on VOI (Campbell et al., 2015; Claxton and Sculpher, 2006; Jalal et al., 2015; Jalal and Alarid-Escudero, 2017). However, for the purposes of this study we sidestep both structural and estimation errors because we specify an underlying event generation process in which the structure and values of the parameters are pre-specified.

Even when a parameter’s “true” value is known, the realisation and timing of events among otherwise identical patients may vary. This stochastic (first-order) variation is inherent to modeling types that utilize Monte Carlo methods to simulate individual patient trajectories. For these methods, a series of simulations, each with fixed simulated patient size *M*, will produce varying estimates of model outputs. Absent any other errors, this series of simulations will yield a set of estimates that center around the outcome’s expected value—and as *M* increases this set of estimates will converge towards this value. But as long as *M* is finite, any given model simulation result will deviate from the expected value. Mirroring the literature, we classify this type of error as **stochastic error** (Briggs et al., 2012). Cohort STMs and DEQ modeling do not suffer from stochastic error because expected values of model outcomes are deterministic solutions from the model.

Stochastic error can never be eliminated (doing so would require simulating an infinite population) but it can be reduced. Best practice recommends a sufficient number of Monte Carlo draw’s to reduce stochastic error to within an acceptable tolerance level (Briggs et al., 2012)—though as we show below, the number of simulated patients needed to achieve a fixed tolerance level is significantly higher with microsimulation than with DES.

Models that coarsen the timing of events into discrete cycles (e.g., cohort-based Markov models and discrete-time and -state microsimulation models) can exhibit additional sources of error. **Integration error** occurs when the model accumulates events at time cycle boundaries (either at the beginning or the end) rather than at any point in continuous time. Depending on whether events accumulate at the beginning or the end of the cycle, this results in over- or under-estimation of time spent in the cycle. An important historical literature in applied mathematics dating back to the 1700s has developed approaches to adjust estimates for this type of error (Boole, 1880; Epperson, 2013; Simpson, 1743). Applications in health economics and health technology assessment frequently rely on half-cycle corrections or the life-table method, but other methods (e.g., those based on Simpson’s rule) can achieve an even greater degree of accuracy (Barendregt 2009).

Discrete time models (e.g., MARKOV, MICROSIM) exhibit **embedding error** if transition probabilities are not specified to align with the underlying continuous time process. These errors are most likely to occur in models that rely on literature-based parameters that are transformed and embedded into transition probability matrices using widely-used conversion formulas (e.g., conversion of event rates to probabilities using *P = 1 − e^−rt^*, where *r* is the rate and *t* is the time cycle). These errors are also more likely to occur in circumstances where events are clustered near each other (e.g., time-varying probabilities of complications or events after a procedure, drug initiation, etc.).

Embedding errors arise because if not applied properly, standard conversion formulas do not separately accommodate more than one competing event (Jahn et al., 2019; Jones et al., 2017; O’Mahony et al., 2015; van Rosmalen et al., 2013; Welton and Ades, 2005). This observation is relevant because in many applications, the modeled process is conceptualised around multiple competing events or sequences of events occurring in continuous time (e.g., progression to or among various disease states or to an absorbing death state).

If probabilities are not embedded properly, standard rate-to-probability conversion processes effectively rule out the possibility of two or more events occurring in the same cycle. Suppose, for example, that the underlying process progresses along three states: A→B→C. When specified to approximate the underlying continuous time process, the probabilities in a transition matrix should reflect the possibility of both B and C occurring in the same cycle. That is, from state A there are two event sequences that can happen within a single cycle: (1) A→B; and (2) A→B→C.

Accounting for these event sequences requires an embedded transition matrix that includes a non-zero probability of transition from A to C by the end the cycle—that is, a transition that is seemingly inconsistent with the underlying disease progression process. This need arises because the simple conversion of the rate of *B* (*r_B_* to the probability of 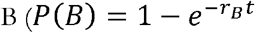) returns the marginal probability of event B occurring within the cycle. This marginal probability is the sum or union of *all* the probabilities of experiencing the event, i.e., P(B) = P(A→B) + P(A→B→C).^ii^ Thus, embedding the marginal probability as the probability of A→B transitions in a cycle will both overstate the correct A→ B transition probability and rule out any A→ B→C transitions within a cycle.

This implicit assumption (i.e., that there are no A→B→C transitions within the time cycle) means that an improperly embedded transition matrix no longer approximates a continuous time process. More technically, it means that the process may no longer be expressed in terms of an underlying generator matrix of continuous transition intensities (Iskandar, 2018). Specifying a transition matrix consistent with an underlying generator matrix is important not only for accurate modeling of the underlying continuous time process, but also for changing Markov cycle lengths (Chhatwal et al., 2016; Jahn et al., 2019). Otherwise, matrix transformation routines (e.g., based on unit roots from eigendecomposition of a specified transition probability matrix) can result in transformed transition matrices with negative probabilities and/or without unique solutions (Chhatwal et al., 2016; Jahn et al., 2019).

Approaches to addressing embedding error complicate the structure and execution of discrete time cohort and microsimulation models. If transitions can first be specified (or estimated) as rates and summarized in a transition intensity matrix, then properly embedding a transition probability matrix is straightforward using modern statistical software: one simply takes the matrix exponential of the transition intensity matrix scaled by the time cycle duration. The result is the solution to Kolmogorov’s forward equation, and this matrix can be used as the transition probability matrix for the model (Jones et al., 2017; Welton and Ades, 2005). Note, however, that this transformation process requires the inclusion of non-Markovian event “accumulators” in the transition matrix. These accumulators ensure that model outputs (e.g., average QALYs and costs) account for events that occur during “jump-over” states that arise from the embedding process (i.e., the model must be structured to account for any cost or utility implications of event B for the A→C transitions in the example above). The appendix and replication code to this study demonstrates how to incorporate non-Markovian accumulators within a transition probability matrix.

One tempting solution to address embedding error is to reduce the cycle length so that the likelihood of multiple transitions within a single cycle approaches zero. Shortening the cycle length carries additional computational costs. More importantly, however, a unique solution to a time step transformation still requires a generator matrix of continuous transition intensities as specified above. Otherwise, converting (marginal) literature-based probabilities to rates, and then using these rates to change the time step (e.g., using eigendecompositions) can result in nonunique solutions and/or even negative probabilities (Chhatwal et al., 2016). Thus, when starting from a transition probability matrix (as opposed to a transition intensity matrix), shortening the time step does not fundamentally address embedding error issues.

Finally, microsimulation models also suffer from **truncation error**, which introduces variance and a small amount of bias to the modeled estimates. This error occurs because information on the precise timing of an event occurring in continuous time is lost when events times are “rounded up” or “rounded down” to the boundary point in the cycle. These rounding errors, when aggregated across individual simulated patients in a microsimulation, distort both the mean and the variance of outcome estimates relative to what would be estimated in a continuous time model (e.g., DES) that can specify each individual’s event timing to any degree of precision (Tricker, 1984).^iii^

Recall from the discussion above that relative to DES, microsimulation models require a greater number of simulated patients to achieve the same tolerance level. This requirement derives, in part, from the increase in variance from truncation error.^iv^ To reduce the influence of truncation error, modelers can increase the number of simulated patients or can reduce the cycle length (e.g., from 1 year to 1 day) to allow for more precise timing of events. Both of these strategies carry additional computational demands, however.

## 2. Methods

### 2.1 Application: Pharmacogenomics

Our modeling application draws on ongoing work in pharmacogenomics (PGx). PGx involves the use of genetic testing to guide drug selection and/or dosing based on associations between genetic phenotypes and drug metabolism. All model parameters and their baseline values are summarized in Table 2. These values were selected to both approximate a common PGx scenario and to produce results that highlight differences across modeling approaches.

**Table 2.** Model Parameters.

| Description | Parameter Name | Parameter Value |
| --- | --- | --- |
| Discount rate | disc | 0.03 |
| DESimulations to perform (1000s) | n | 1000 |
| Diff Eq Time step for DEQ approach | resolution | 0.019 |
| Time horizon (years) of simulation | horizon | 40 |
| Willingness to pay threshold used for NMB (\$1000s / QALY) | wtp | 100 |
| Shape Parameter from Gompertz model of secular death for 40yr female fit using 2012 U.S Social Security mortality data. | shape | 0.101 |
| Rate Parameter from Gompertz model of secular death for 40yr female fit using 2012 U.S Social Security mortality data. | rate | 0.001 |
| Population prevalence of genetic variant. | p_g | 0.2 |
| Condition indication percentage | r_a_pct | 10 |
| Condition indication time period (years) | r_a_dur | 10 |
| Probability of ordering the genetic test. | p_o | 1 |
| Probability of death from adverse drug event. | p_bd | 0.1 |
| Adverse drug event percentage | r_b_pct | 25 |
| Adverse drug event time period (years) | r_b_dur | 1 |
| Relative risk of adverse drug event among patients with genetic variant present and alternative therapy prescribed. | rr_b | 0.8 |
| Initial cost at indication (\$1000s). | c_a | 10 |
| Cost of standard drug therapy. | c_tx | 0.25 |
| Cost of pharmacogenomic alternative drug therapy. | c_alt | 3 |
| Cost of adverse drug event survival (\$1000s). | c_bs | 25 |
| Cost of adverse drug event case fatality (\$1000s). | c_bd | 20 |
| Cost of genetic test. | c_t | 200 |
| Disutility of developing condition. | d_a | 0.05 |
| Disutility duration (years) after developing condition. | d_at | 1 |
| Lifetime disutility of adverse drug event among survivors. | d_b | 0.15 |
| DES= Discrete Event Simulation |  |  |
| NMB = Net Monetary Benefit |  |  |
| QALY = Quality-adjusted life years |  |  |

We focus on a simple and common PGx decision problem: whether to utilize testing to guide drug selection for individuals with a genetic variant that can affect metabolism of a chronic disease medication. Specifically, we consider a population of healthy 40-year-old women at risk of developing an indication (10% incidence rate over 10 years) for a health condition with a standard maintenance medication therapy, but where there is a more expensive and more effective pharmacogenomic alternative available to individuals with a variant.

We assume all who develop the chronic condition incur a one-time treatment cost of $10,000, a transient (one-year) 0.05 utility decrement, and are placed on daily maintenance medication ($0.25 per day) for life. Individuals on this medication are at risk of an adverse event that affects 25% of individuals in the first year they are on the drug. The adverse event has a 10%) case fatality rate, carries a one-time $20,000 cost among decedents and, among the survivors, a one-time $25,000 cost and 0.1 utility decrement for life.

We compare the above reference case (“no testing”) scenario to an alternative scenario (“PGx”) in which all individuals who develop the condition receive a ($200) genetic test as part of their initial diagnosis. Patients who test positive for the variant (20%) population prevalence) are placed on a more expensive ($3/day) alternative maintenance medication for the remainder of their life. This medication lowers the risk of the adverse event (relative risk 0.8). Individuals without the variant do not benefit from taking the pharmacogenomic alternative (relative risk 1.0), so remain on the standard ($0.25/day) therapy.

We model a 40-year time horizon with background mortality based on age- and gender-based mortality as summarized using a Gompertz model fit to U.S. life-table data (Graves et al., 2018). We assumed that all events were not recurrent. All costs are provided in $2019. We also assume a standard discount rate of 0.03 for both health state utilities and costs. Key model outputs included average discounted QALYs and costs, the incremental cost-effectiveness ratio (ICER) comparing the PGx vs. no testing strategies, and Net Monetary Benefit (NMB) based on a willingness-to-pay (WTP) of $100,000 per QALY.

### 2.2 Simulation Models

We developed four simulation models: (1) a differential equations |DEQ| model, which we used as the reference model for comparison with other models since it deterministically solved for average cost and QALY outcomes; (2) a cohort-based discrete-time state transition model [MARKOV]; (3) an individual discrete time state transition model [MICROSIM]; and (4) a discrete event simulation model [DES]. For our primary results the MARKOV and MICROSIM models utilized 10 million simulated patients. For the MARKOV and MICROSIM models we also produced results based on three cycle lengths: a one-year cycle (1Y), a monthly (1M) cycle, and (where feasible) a daily cycle (1D).

For MICROSIM and MARKOV, we modeled each model-cycle-length combination using two approaches: (1) by converting rates to probabilities using a standard conversion formula, i.e., *P*(*t*) = 1 −*e^−rt^*, where *r* is the rate and *t* is the time cycle; and (2) by embedding transition probabilities by taking the matrix exponential of the transition intensity matrix **(G)** as defined by the rates as summarized above and in Table 2, i.e., **P(t) *e^tG^***. We augmented the properly embedded models with non-Markovian accumulators to ensure that utility and cost estimates accounted for competing within-cycle events. We label results based on this embedded transition intensity matrix using -EMB in the model output.

### 2.3 Stochastic Convergence and Model Run Time

We investigated stochastic convergence of estimates of the NMB for the DES and MICROSIM models as compared with deterministic NMB estimates from the DEQ and MARKOV-based models. We did so by varying the number of Monte Carlo draws from one thousand to 1 billion patients, though we also considered Monte Carlo simulation sizes up to 100 billion patients using a bootstrapping procedure that drew (with replacement) from the 1 billion originally simulated patients.

We assessed model run time using a benchmarking procedure that re-estimated each model 100 times using different random seeds. From these results we measured key moments (mean, variance) in the run-time distribution. For this benchmarking procedure we simulated ten million patients for the DES and MICROSIM models. To ensure fair comparison, runtime benchmarking was run on a high-performance computing cluster requesting 1 CPU and 120G memory.

Reproducible model code is included in the supplemental material to this manuscript.

## 3. Results

### 3.1 Baseline Results

Based on the DEQ model, PGx testing yielded an ICER of $103,212/QALY vs. a reference no-testing strategy (Table 3). A Markov model with an annual cycle duration and transition probabilities converted using the standard formula (P(t)= 1 − e^−rt^) resulted in an ICER of $91,929/QALY. Therefore, at a willingness-to-pay threshold of $100,000/QALY, different decisions on the optimal treatment strategy would be made depending on the choice of a Markov (annual cycle and standard probability conversion) vs. DEQ model. Only at successively shorter cycle durations (e.g., 1 month and 1 day with standard probability conversion) did the Markov model results begin to resemble those obtained using DEQ. For example, a Markov model with a daily time cycle yielded an ICER of $103,173/QALY.

**Table 3.**
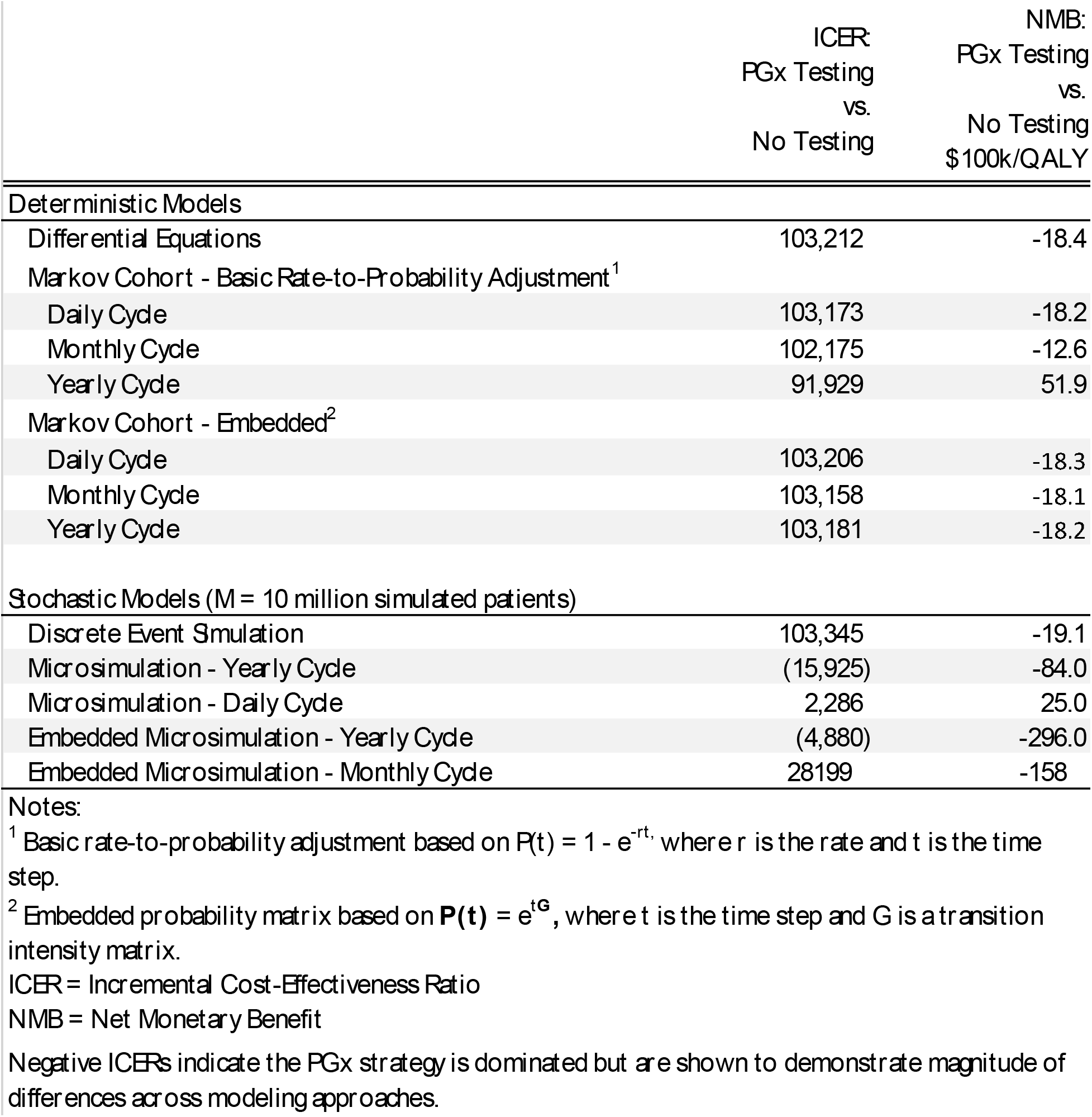
Outcomes by Model Type

By contrast, cohort state transition models embedded by exponentiating the transition intensity matrix yielded ICERs comparable to the DEQ for all time cycle durations (Table 3). For example, with a yearly time cycle the properly embedded Markov produced an ICER of $103,181 vs. $103,212 using DEQ.

Among stochastic models with 10 million simulated patients, only DES yielded ICER and NMB results similar to the DEQ and MARKOV-EMB models. The DES produced an ICER of $103,345 while the microsimulation models—including those that used embedded transition probability matrices—yielded ICERs ranging from -$15,925 (i.e., PGx strategy dominated) to $28,199.

### 3.2 Stochastic Model Convergence

Among the Monte Carlo-based models considered, convergence with deterministic outcomes from the DEQ was achieved with considerably fewer simulated patients for DES as compared with MICROSIM (Figure 1). Only with short (1-day) cycle durations and/or embedding corrections and 1 billion or more simulated patients did the MICROSIM models consistently estimate similar outcomes as the DEQ or MRKCIIT-EMB models. By contrast, DES models with 1 million patients or more reliably produced average NMB outcomes similar to the DEQ and MARKOV-EMB models.

**Figure 1.**
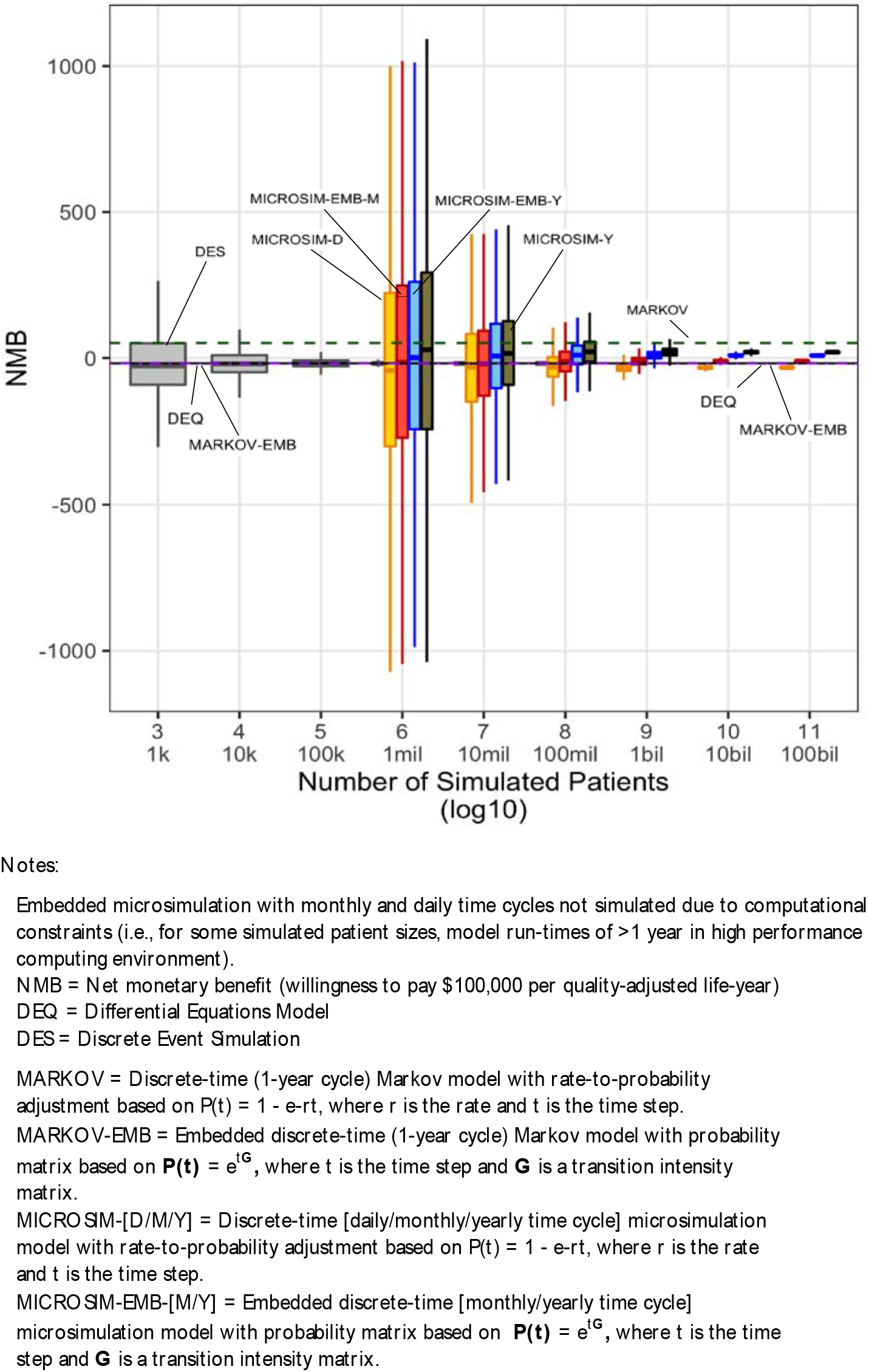
Convergence of Net Monetary Benefit Outcome for Stochastic Models to Deterministic Model Outputs.

### 3.3 Computation ïime

Computation time differed across modeling approaches. Run-time was fastest for deterministic solutions based on DEQ (median runtime 3.6sec) and MARKOV with a yearly cycle (median=0.01 sec using standard conversion formulas and 1.1 sec for the embedded model based on the transition intensity matrix). By comparison, a Markov model with daily cycle duration had a median runtime of 8.7 minutes. Among Monte-Carlo-based models, the DES had a median run time of 48.2 seconds for 10 million patients, while MICROSIM with an annual cycle duration had a median run-time of 19.0 minutes. Due to computational constraints we were unable to test the run-time for a MICROSIM with a daily cycle length.^v^

**Figure 2.**
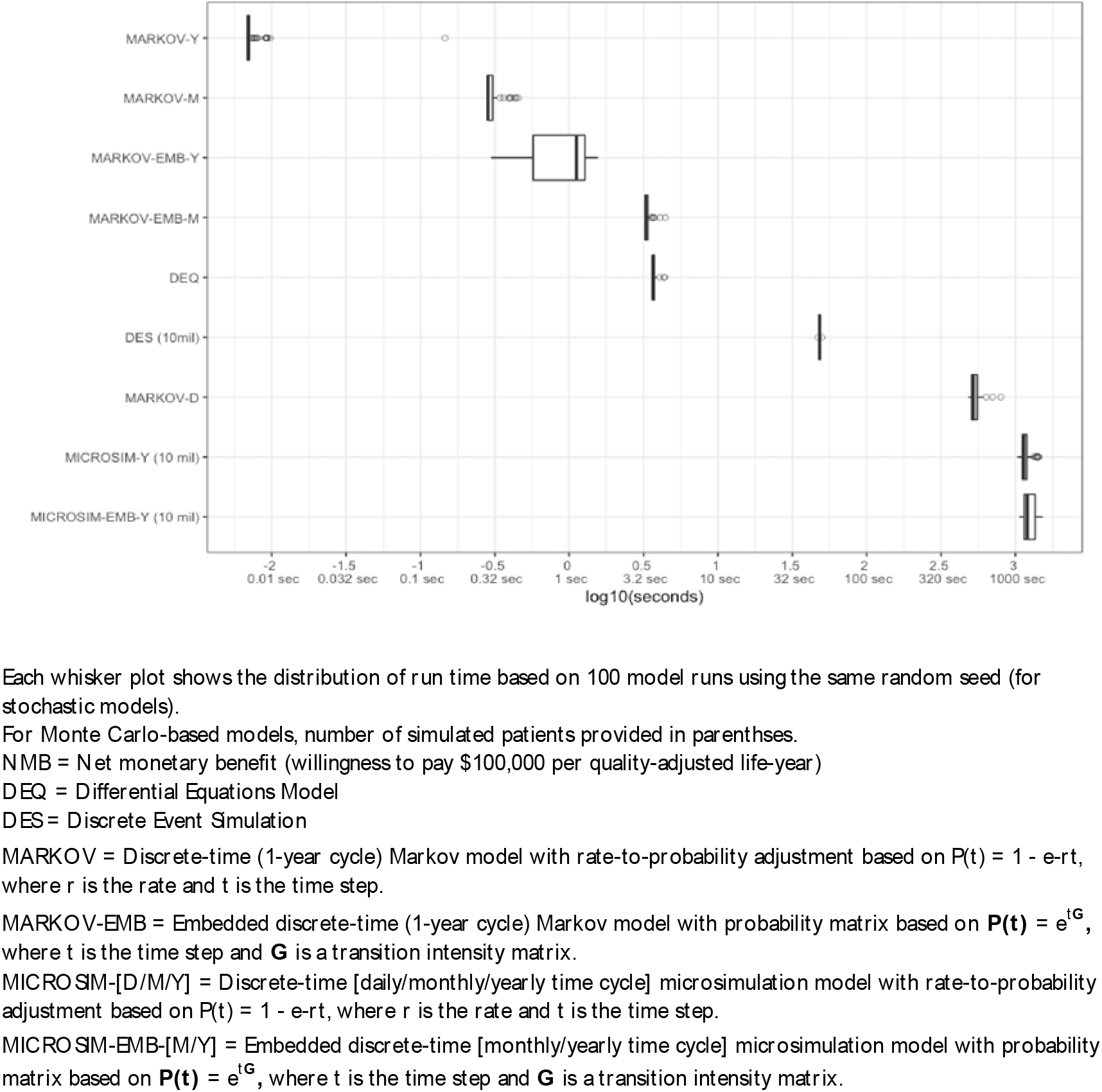
Model Run Time Comparison

## 4. Discussion

This study demonstrates that modeling choices affect the scope, accuracy and reliability of decision outcomes in health economic assessments. By comparing outputs and optimal decisions across four commonly used modeling approaches, we show that care must be taken when structuring and executing models.

Given the predominance of discrete time cohort state transition (Markov) models in health technology assessments (Karnon, 2003), one noteworthy observation from our study is that embedding transitionmatrices using commonly-used rate-to-probability conversion formulas can de-couple a Markov model from the continuous time process it aims to represent. In essence, an improperly-embedded discrete time model will rule out events and event sequences from occurring within a time step that would, with some probability, occur in continuous time. While this is certainly not a new observation (Jahn et al., 2019; Jones et al., 2017; O’Mahony et al., 2015; van Rosmalen et al., 2013; Welton and Ades, 2005), our results demonstrate that improper embedding can meaningfully affect decision outcomes.

Our results also highlight a telltale sign of embedding problems: outcomes change as shorter time cycles are used. While it may be tempting to simply adopt a shorter time cycle to reduce the likelihood of two competing events occurring within the time step, it is worth emphasizing that a unique solution to time-step transformations requires that the transition matrix be expressible in terms of a generator matrix of transition intensities—precisely the matrix needed to properly embed the original transition matrix in the first place (Chhatwal et al., 2016). In other words, simple transformations of rates to probabilities for a shorter time cycle does not guarantee a unique or even correct solution.

Our study was also the first to run a head-to-head “horse-race” between Monte Carlo-based modeling approaches. In this case, DES demonstrated clear advantages. DES was both computationally more efficient and resulted in decision outcomes that were nearly identical (with a sufficient number of simulated patients) to DEQ and embedded Markov models. By contrast, microsimulation required substantially more simulated patients to yield reliable results and, when it did converge, resulted in slightly different average outcomes (though no difference in optimal decisions) even when using embedded transition matrices.

One potential reason for this nonconvergence merits additional discussion because it highlights an additional limitation of microsimulation. In our model application, the adverse event occurs downstream of the drug indication. Thus, the probability of both drug indication *and* an adverse event both occurring within a yearly (or even monthly) time cycle is non-zero. A properly embedded transition matrix would include both a probability of transitioning from “healthy” to “indication,” as well as a probability of transitioning from “healthy” to “adverse event” (with non-Markovian accumulators added to capture the sojourn through “indication”). Critically, however, these embedded probabilities change depending on the decision to place the patient on the alternative drug. For example, the probability of a transition from “healthy” to “adverse event” within a time step is smaller if the alternative drug was used, since the risk of an adverse event is lower in that case. Because the overall (marginal) probability of drug indication is fixed (PGx only affects which drug is prescribed, not the drug indication itself), this would result in a higher probability transition from “healthy” to “indication” if the patient received the alternative drug. In other words, the initial probability of transition from “healthy” to “indication” changes depending on downstream decisions in the model. This cart-and-horse problem complicates the structure and execution of microsimulation because it requires that all potential decisions be modeled *before* a patient can transition through the model. To address it, a microsimulation model must either first solve a Markov model to determine the correct probabilities, or must be structured more similarly to a DES.

To conclude, it is worth offering guidance based on our study that can aid future modeling decisions for other researchers. For standard health economic assessments, Markov models offer good balance between speed and accuracy. However, if competing events or event sequences are clustered near each other (e.g., complications after a procedure or prescription), modelers should take great care to ensure that embedding biases do not materially affect decision outcomes. Proper embedding complicates model structure since non-Markovian accumulators must be included to capture the cost and utility changes from sojourns implied by “jumpover” states. However, a correctly embedded transition matrix also means that model results will not change if longer time cycles are used. This could be important in applications that aim to utilize computationally intensive probabilistic sensitivity analysis (PSA) and VOI methods to understand model sensitivity and guide future work (e.g., the expected value of sample information). That is, rather than a (computationally demanding) monthly cycle, a yearly cycle could be used to conduct VOI analyses based on a properly embedded Markov model. Finally, our results make clear that if the modeling scenario calls for the use of a Monte-Carlo model, DES models avoid many of the pitfalls that complicate microsimulation and yield a favorable mix of accuracy, reliability, and speed.

## Data Availability

All replication code and data for this manuscript is available as part of the supplementary material. Please contact the lead author (Graves) for access.

## Footnotes

i We are grateful for feedback on this study from participants at the 40^th^ Annual North American Research Meeting of the Society for Medical Decision Making. Financial support for this study was provided by the NIH Common Fund (U01HL122904) and the National Human Genome Research Institute (1R01HG009694-01). The funding agreement ensured the authors’ independence in designing the study, interpreting the data, writing, and publishing the report. We also thank Ashley Leech for her thoughtful review of our manuscript. The authors have no conflicts to report.

ii The same logic still holds if events B and C were simply competing and not sequential events, in which case the marginal probability of event B reflects P(A→B) and P(A→B & A→C).

iii Intuitively, integration error occurs when events are *moved* from their precise time to the beginning or end of the cycle. Truncation error arises because information on the precise timing of events is lost when all events in the cycle are truncated to the cycle boundary point.

iv In the Appendix, we demonstrate how truncation error can affect model output variance by constructing a straw-man DES model that truncates DES event times at a time interval (year). Because DES models model the precise timing of events, a comparison of a standard DES to a truncated DES model (holding all other parameters and random seeds fixed) can isolate the contribution of truncation error. The plot of model estimate convergence shows clearly that the variance of model outputs is considerably greater with the truncated DES model.

v To provide context for how long a daily MICROSIM would take to complete, a cohort-based Markov model with a daily cycle had a median 522sec of runtime, as compared with 0.00697sec for the same model with an annual cycle length—nearly 75,000 times longer. Using this ratio to scale the yearly MICROSIM result (19.0 minutes) implies a model run time of 23,758 hours.

## References

Baltussen, R., Marsh, K., Thokala, P., Diaby, V., Castro, H., Cleemput, I., Garau, M., Iskrov, G., Olyaeemanesh, A., Mirelman, A., Mobinisadeh, M., Morton, A., Tringali, M., Til, J. van Valentim, J., Wagner, M., Youngkong, S., Zah, V., Toll, A., Jansen, M., Bijlmakers, L., Oortwijn, W., Broekhuisen, H., 2019. Multicriteria Decision Analysis to Support IITA Agencies: Benefits, Limitations, and the Way Forward. Value in Health 0. https://doi.org/10.1016/j.jval.2019.06.014

Baltussen, R., Niessen, L., 2006. Priority setting of health interventions: the need for multi-criteria decision analysis. Cost effectiveness and resource allocation 4, 14.

Barendregt, J.J., 2009. The half-cycle correction: banish rather than explain it. Medical Decision Making 29, 500–502.

Brennan, A, Chick, S.E., Davies, R., 2006. A taxonomy of model structures for economic evaluation of health technologies. Health economics 15, 1295–1310.

Briggs, A., 2006. Decision Modelling for Health Economic Evaluation, 1 edition, ed. Oxford University Press, USA, Oxford.

Briggs, A.H., Weinstein, M.C., Fenwick, E.A., Karnon, J., Sculpher, M.J., Paltiel, A.D., 2012. Model parameter estimation and uncertainty analysis: a report of the ISPOR-SMDM Modeling Good Research Practices Task Force Working Group–6. Medical decision making 32, 722–732.

Campbell, J.D., McQueen, R.B., Libby, A.M., Spackman, D.E., Carlson, }.}., Briggs, A, 2015. Cost-effectiveness uncertainty analysis methods: a comparison of one-way sensitivity, analysis of covariance, and expected value of partial perfect information. Medical Decision Making 35, 596–607.

Caro, J.J., 2016. Discretely integrated condition event (DICE) simulation for pharmacoeconomics. Pharmacoeconomics 34, 665–672.

Caro, J.J., Möller, J., 2016. Advantages and disadvantages of discrete-event simulation for health economic analyses. Expert Review of Pharmacoeconomics & Outcomes Research 16, 327–329. https://doi.org/10.1586/14737167.2016.1165608

Chhatwal, J., Jayasuriya, S., Elbasha, E.H., 2016. Changing cycle lengths in state-transition models: challenges and solutions. Medical Decision Making 36, 952–964.

Claxton, K.P., Sculpher, M.J., 2006. Using value of information analysis to prioritise health research. Pharmacoeconomics 24, 1055–1068.

Drummond, M.F., Sculpher, M.J., Claxton, K., Stoddart, G.L., Torrance, G.W., 2015. Methods for the economic evaluation of health care programmes. Oxford university press.

Epperson, J.F., 2013. An introduction to numerical methods and analysis. John Wiley & Sons.

Graves, J.A., Garbett, S., Zhou, Z., Peterson, J., 2018. The Value of Pharmacogenomic Information.

Heath, A, Kunst, N.R., Jackson, C., Strong, M., Alarid-Escudero, F., Goldhaber-Fiebert, J.D., Baio, G., Menzies, N.A., Jalal, H., 2019. Calculating the Expected Value of Sample Information in Practice: Considerations from Three Case Studies. arXiv preprint arXiv:1905.12013.

Iskandar, R., 2018. A theoretical foundation for state-transition cohort models in health decision analysis. PLOS ONE 13, e0205543. https://doi.org/10.1371/journal.pone.0205543

Jacobson, S.H., Hall, S.N., Swisher, J.R., 2006. Discrete-event simulation of health care systems, in: Patient Flow: Reducing Delay in Healthcare Delivery. Springer, pp. 211–252.

Jahn, B., Kursthaler, C., Chhatwal, J., Elbasha, E.H., Conrads-Frank, A, Rochau, U., Sroczynski, G., Urach, C., Bundo, M., Popper, N., 2019. Alternative Conversion Methods for Transition Probabilities in State-Transition Models: Validity and Impact on Comparative Effectiveness and Cost-Effectiveness. Medical Decision Making 0272989X19851095.

Jalal, H., Alarid-Escudero, F., 2017. A Gaussian Approximation Approach for Value of Information Analysis. Medical Decision Making 0272989X1771562. https://doi.org/10.1177/0272989X17715627

Jalal, H., Goldhaber-Fiebert, J.D., Kuntz, K.M., 2015. Computing expected value of partial sample information from probabilistic sensitivity analysis using linear regression metamodeling. Medical Decision Making 35, 584–595.

Jones, E., Epstein, D., García-Mochón, L., 2017. A procedure for deriving formulas to convert transition rates to probabilities for multistate Markov models. Medical Decision Making 37, 779–789.

Karnon, J., 2003. Alternative decision modelling techniques for the evaluation of health care technologies: Markov processes versus discrete event simulation. Health economics 12, 837–848.

Karnon, J., Stahl, J., Brennan, A., Caro, J.J., Mar, J., Möller, J., 2012. Modeling using discrete event simulation: a report of the ISPOR-SMDM Modeling Good Research Practices Task Force–4. Medical decision making 32, 701–711.

Kunst, N.R., Wilson, E., Alarid-Escudero, F., Baio, G., Brennan, A., Fairley, M., Glynn, D., Goldhaber-Fiebert, J.D., Jackson, C., Jalal, H., 2019. Computing the Expected Value of Sample Information Efficiently: Expertise and Skills Required for Four Model-Based Methods. arXiv preprint arXiv:1910.03368.

Neumann, P.J., Sanders, G.D., Russell, L.B., Siegel, J.E., Ganiats, T.G., 2016. Cost-effectiveness in health and medicine. Oxford University Press.

O’Mahony, J.F., Newall, A.T., van Rosmalen, J., 2015. Dealing with Time in Health Economic Evaluation: Methodological Issues and Recommendations for Practice. Pharmacoeconomics 33, 1255–1268. https://doi.org/10.1007/s40273-015-0309-4

Peacock, S., Mitton, C., Bate, A., McCoy, B., Donaldson, C., 2009. Overcoming barriers to priority setting using interdisciplinary methods. Health Policy 92, 124–132. https://doi.Org/10.1016/j.healthpol.2009.02.006

Rutter, C.M., Zaslavsky, A.M., Feuer, E.J., 2011. Dynamic Microsimulation Models for Health Outcomes: A Review. Med Decis Making 31, 10–18. https://doi.org/10.1177/0272989X10369005

Siebert, U., Alagoz, O., Bayoumi, AM., Jahn, B., Owens, D.K., Cohen, D.J., Kuntz, K.M., 2012. State - Transition Modeling: A Report of the ISPOR-SMDM Modeling Good Research Practices Task Force–3. Med Decis Making 32, 690–700. https://doi.org/10.1177/0272989X12455463

Simpson, T., 1743. Mathematical Dissertations on a Variety of Physical and Analytical Subjects. Containing, Among Other Particulars, a Demonstration of the True Figure which the Earts,… A General Investigation of the Attraction at the Surfaces of Bodies Nearly Sphrical… The Whole in a General and Perspicuous Manner. By Thomas Simpson. T. Woodward, at the Half-Moon, between the two Temple-Gates in Fleetstreet.

Stahl, J.E., 2008. Modelling methods for pharmacoeconomics and health technology assessment. Pharmacoeconomics 26, 131–148.

Standfield, L., Comans, T., Scuffham, P., 2014. Markov modeling and discrete event simulation in health care: a systematic comparison. International journal of technology assessment in health care 30, 165–172.

Thokala, P., Duenas, A, 2012. Multiple criteria decision analysis for health technology assessment. Value in Health 15, 1172–1181.

Tricker, A.R., 1984. Effects of rounding on the moments of a probability distribution, journal of the Royal Statistical Society: Series D (The Statistician) 33, 381–390.

Welton, N.J., Ades, A.E., 2005. Estimation of Markov chain transition probabilities and rates from fully and partially observed data: uncertainty propagation, evidence synthesis, and model calibration. Medical Decision Making 25, 633–645.

